# Ultrasensitive measurement of both SARS-CoV2 RNA and serology from saliva

**DOI:** 10.1101/2021.01.25.21249679

**Authors:** Dmitry Ter-Ovanesyan, Tal Gilboa, Roey Lazarovits, Alexandra Rosenthal, Xu G. Yu, Jonathan Z. Li, George M. Church, David R. Walt

## Abstract

Tests for COVID-19 generally measure SARS-CoV2 viral RNA from nasal swabs or antibodies against the virus from blood. It has been shown, however, that both viral particles and antibodies against those particles are present in saliva, which is more accessible than both swabs and blood. We present methods for highly sensitive measurements of both viral RNA and serology from the same saliva sample. We developed an efficient saliva RNA extraction method and combined it with an ultrasensitive serology test based on Single Molecule Array (Simoa) technology. We apply our test to the saliva of patients who presented to the hospital with COVID-19 symptoms, some of whom tested positive with a conventional RT-qPCR nasopharyngeal swab test. We demonstrate that combining viral RNA detection by RT-qPCR with serology identifies more patients as infected than either method alone. Our results suggest the utility of combining viral RNA and serology testing from saliva, a single easily accessible biofluid.

## Introduction

The two main tests for SARS-CoV2 infection are molecular tests to detect the presence of the virus (RNA or antigen) and serological tests to detect the presence of antibodies against the virus^1–6^. Both tests have advantages and disadvantages. RT-qPCR, the main diagnostic test and current gold standard, is a sensitive method for measuring the presence of viral RNA, usually performed from nasopharyngeal (NP) or anterior nasal swabs^1–3^. However, it has been shown that not all patients who are infected with SARS-CoV2 test positive for viral RNA^1,2,5,7–9^. There are several potential reasons for this: low viral load, variability in swabbing, or late swab collection relative to the time of infection^2,5^. The time at which the swab is performed is important because, after initial infection, levels of virus sharply rise and then drop, providing a relatively narrow window at which viral RNA is present^1,5,6,10–18^. Serology tests detect antibodies that develop against the virus during infection^1,6^. These antibodies remain stable for at least several months, widening the time window of testing for SARS-CoV2 infection^19–21^. Combining RNA detection with serology testing has the potential to increase the sensitivity of RT-qPCR alone^6,12,20,22–26^ by reducing false negatives in RT-qPCR.

RT-qPCR tests for SARS-CoV2 mostly analyze RNA from nasal swabs, while serology tests are generally performed using blood^1,22,27–29^. However, studies have shown that both SARS-CoV2 viral RNA^12,30–33^ and antibodies^19,30,34^ against the virus are present in saliva. Since saliva is easier to collect than either swabs or blood, its accessibility makes it an ideal biofluid for widespread diagnostic use. Research is still ongoing regarding how well saliva correlates to different types of swabs in terms of sensitivity of viral RNA detection^30–32,35,36^. This will depend largely on several factors such as patient selection, sample collection, and RNA extraction methodology. Nonetheless, it is clear that nSARS-CoV2 viral RNA can be readily detected in saliva^12,30–32^. Similarly, previous studies have shown a strong correlation between antibodies against SARS-CoV2 in blood and saliva^19,30,34,37–39^.

Although RT-qPCR is highly sensitive, there is great variability in the RNA extraction efficiency, which is generally performed using commercial kits. These kits have advantages in terms of ease, but are prone to supply chain limitations^40^. Furthermore, using kits may lead to incompatibilities with upstream or downstream steps since the components are unknown to the user. Lastly, kits are not optimized for specific biofluids such as saliva, leading to potentially low RNA recovery. Serology testing in saliva also presents challenges; namely, that concentrations of antibodies in saliva are much lower than they are in blood^19,30,34^ and that different isotypes may be present in the different fluids.

We set out to develop a highly sensitive test to detect both viral RNA and antibodies against the virus from the same saliva sample. We developed an optimized RNA extraction protocol that is highly efficient for saliva. We also adapted a Single Molecule Array^41^ (Simoa)-based ultrasensitive serology test we previously developed for detecting SARS-CoV2 antibodies in blood for use in saliva^7,42^. Combining our RNA extraction protocol for RT-qPCR with this ultrasensitive serology test, we are able to better classify COVID-19 patients as being actively or previously infected with SARS-CoV2, demonstrating the utility of this approach for accurate diagnosis of COVID19.

## Results

In order to develop a highly sensitive test for both SARS-CoV2 RNA and serology using saliva (schematically presented in Figure 1), we first optimized the RNA extraction. We developed two versions: a low-volume version (30μL saliva) that can be performed in a 96-well PCR plate and a high-volume version (300μL saliva) that can be performed in a deep-well plate. We started with a general method based on binding nucleic acids to carboxylated paramagnetic beads in the presence of a guanidinium thiocyanate lysis buffer (Figure 2a), and optimized each step of the protocol using saliva samples. To evaluate our extraction method, we spiked in known amounts of synthetic SARS-CoV2 RNA or heat-inactivated SARS-CoV2 viral particles into PBS or human saliva and then quantified recovery after RNA extraction by RT-qPCR. (Figure 2a, Supplementary Figure 1). We tested a large number of parameters using this approach until we arrived at a protocol that consistently resulted in 50-100% recovery in 30μL of saliva (Figure 2b). To maximize sensitivity, we opted to use more saliva (300μL) in a deep-well plate format. However, as it is difficult to elute into a small volume using a deep well plate, we included a transfer step in our protocol where we transfer the beads (during a 70% ethanol wash step) from a deep-well plate to a PCR plate, allowing us to elute the RNA in only 10μL (Figure 2a).

**Figure 1:**
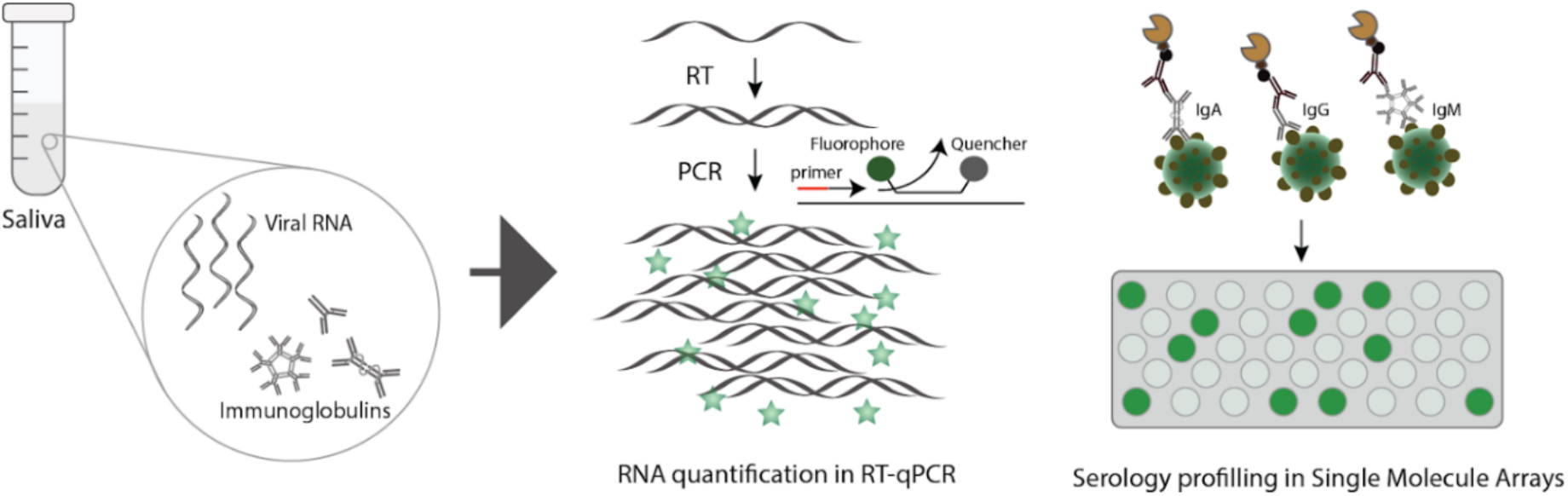
Schematic illustration of the approach for simultaneous detection of SARS-CoV2 RNA and serology in saliva samples. Saliva samples, containing both viral RNA and immunoglobulins, are split for multi-analyte detection. RNA is extracted with our custom protocol and detected using RT-qPCR. Low levels of antibodies are detected using ultrasensitive Simoa assays.

**Figure 2:**
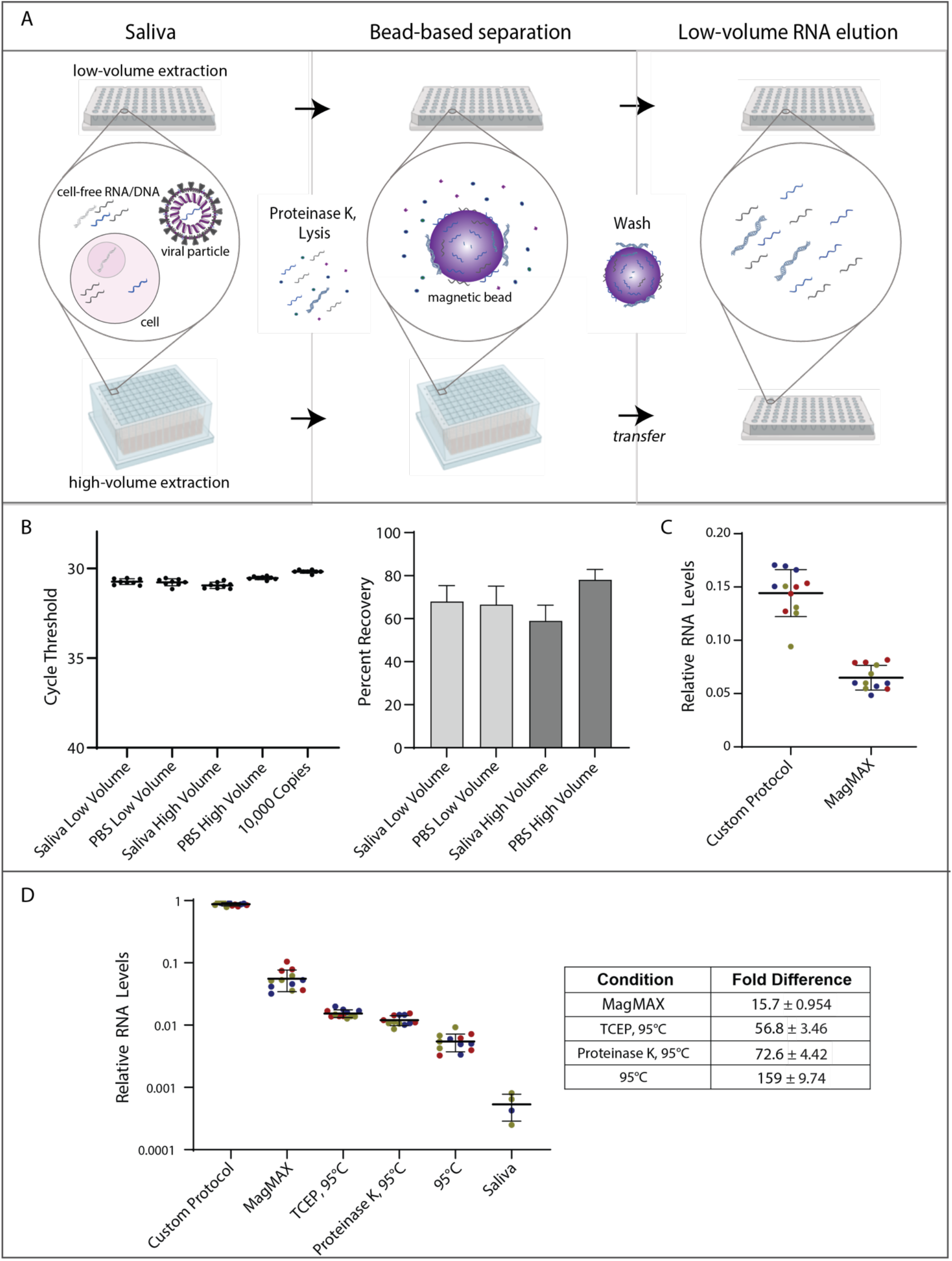
Custom RNA extraction protocol in saliva recovers viral RNA with high efficiency. A) Schematic representation showing our bead-based extraction protocol in low-volumes and high-volumes. B) High efficiency viral RNA recovery from PBS and saliva using custom protocol. PBS or saliva (30μL for low volume or 300μL for high volume) were spiked with 10,000 copies of synthetic SARS-CoV2 RNA (RNA). Our custom RNA extraction protocol was performed, and extracted RNA was compared to equivalent input amounts of spiked in RNA using RT-qPCR. Recovery of RNA is calculated as RNA level of extracted SARS-CoV2 RNA relative to spike in amount. C) Direct comparison of custom protocol with commercial kit using same volume. RNA was extracted from 200μL of saliva with 50 particles/μL of heat-inactivated virus and eluted into 50μL using either our custom protocol or the MagMAX kit and quantified by RT-qPCR. D) Comparison of extraction and extraction-free methods. For all methods, saliva spiked with 50 viral particles/μL was used and the same volume of inactivated saliva or eluted RNA was compared by RT-qPCR. For comparison of RNA extraction methods, maximum input volume (400μL for MagMAX vs. 300μL for custom) and minimal elution volume (50μL for MagMAX vs. 10μL for custom) were used. Relative RNA Levels (log scale) represent RT-qPCR quantification for each condition relative to spike-in aliquot on same RT-qPCR plate. Fold-differences for Relative RNA Levels measured in the Custom Protocol relative to the other conditions are summarized in the table. For the saliva control (no-inactivation), 8/12 replicates had no detectable levels of RNA and were not plotted. Four replicates were repeated across three different days for each condition (different colors represent different days). Error bars for all figures indicate Standard Deviation.

To see how our protocol compares to a commonly used commercial kit, we performed a head-to-head comparison with the ThermoFisher Scientific MagMAX Viral/Pathogen Nucleic Acid Isolation kit. We first extracted RNA from equal volumes of saliva spiked with viral particles and eluted the captured RNA into equal volumes (Figure 2c). In this comparison, we found that our custom protocol had more than twice the recovery of MagMAX. We also compared our protocol to MagMAX using the maximum input volume and minimum elution volumes for both protocols using saliva with the same concentration of viral particles. In this case, our protocol recovered 16x more RNA (Figure 2d). We then compared our protocol to three recently developed protocols for measuring RNA from saliva without purification. These protocols, SalivaDirect^43^, TCEP inactivation^44^, and 95°C heating^45^, all use a small volume of saliva (<10μL) since the final volume of RT-qPCR reactions is usually 20μL. We compared our high-volume RNA extraction to these three protocols using saliva containing the same concentration of viral particles. We found that we detected significantly more RNA in extraction-based protocols, largely due to the ability to use larger volumes of saliva (Figure 2d).

After validation of our RNA extraction protocol, we turned to combining RNA detection with serology in clinical saliva samples from COVID-19 patients. We have previously developed Single Molecule Array (Simoa) based ultrasensitive profiling of IgG, IgM, and IgA antibodies against SARS-CoV2 nucleocapsid (N), spike (S), S1, and RBD protein targets in blood^7^. Our assay employs a bead-based, digital ELISA for high-throughput, automated ultrasensitive detection of antibodies in small volumes. Using only 40μL of saliva per sample (10μL for serology and 30μL for RNA), we characterized 12 antibody interactions and quantified SARS-CoV2 RNA across 18 saliva samples. We tested two pre-pandemic saliva samples from healthy individuals, and 16 samples from symptomatic individuals who visited the MGH Respiratory Infection Clinic during the pandemic (see SI Table S1 for clinical characteristics summary). The patients were all tested by RT-qPCR from NP swab samples upon arrival to the clinic and saliva was collected on the same day.

Combined measurement of SARS-CoV2 RNA and IgG, IgM and IgA levels against S1 (Figure 3a) revealed that five patients were positive for either RNA, serology, or both, in saliva compared to three patients by NP RT-qPCR alone. Two patients with positive serology in saliva but negative RT-qPCR (in both NP swabs and saliva) displayed severe respiratory illness, and thus likely received false negative RT-qPCR results. RNA was detected in saliva for two of the three patients with positive NP swabs. For the patient with RNA detected in the NP swab but not saliva, antibody levels were above the threshold for IgA and IgM against S1. Since our previous work showed that IgA − S1 displayed the best separation between positives and controls, this sample would be classified as positive in our serological assay. We also measured saliva from a subset of the patients at six late time points (>9 days after the first positive PCR). We found no RNA was detected but antibody levels were high for most of the immunoglobulin subtypes we measured, as expected (Figure 3b). Finally, to see if we could achieve multiplexed detection of viral RNA and serology on the same platform from one sample, we developed a Simoa assay for the direct detection of SARS-CoV2 RNA. Although we found this assay to be less sensitive than RT-qPCR, the Simoa assay detects RNA without amplification. We combined this assay with our serology assay to detect RNA and antibodies spiked into saliva samples (SI Figure S3, Table S2), demonstrating multiplexed detection of RNA and serology on the Simoa platform.

**Figure 3:**
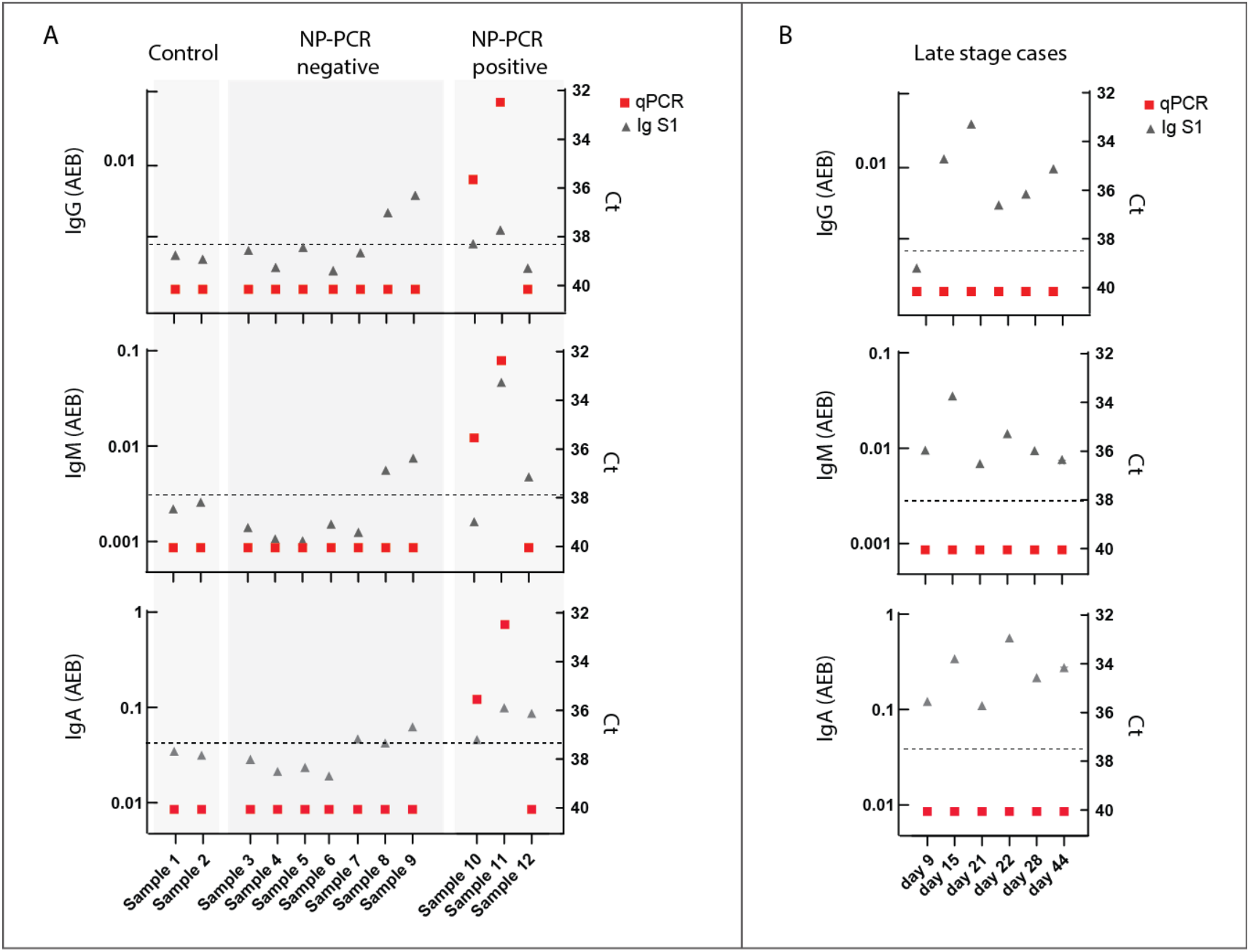
Detection of SARS-CoV2 RNA and serology from the same saliva sample of hospitalized COVID-19 patients. A) Detection of viral RNA and serology from saliva patient samples on day of hospitalization. RT-qPCR Ct levels (red rectangular) and Simoa serological mean AEB (average enzyme per bead) levels (gray triangles) for IgG, IgM, and IgA against S1 subunit. The samples were divided into three groups: pre-pandemic control samples (left, n=2), saliva samples from patients who tested negative by RT-qPCR from nasopharyngeal swabs (NP-PCR negative, middle, n=7) and saliva samples from patients who tested positive by RT-qPCR from nasopharyngeal swabs (NP-PCR positive, right, n=3). Black dotted lines indicate the antibodies threshold above the control samples. Conditions were signal is undetected are set to Ct = 40. B) Detection of viral RNA and serology from saliva patient samples several days (between 9 and 44) after hospitalization. Conditions were signal is undetected are set to Ct = 40.

## Discussion

SARS-CoV2 infection is currently diagnosed with RT-qPCR from NP swabs and the immune response is monitored using serology tests from blood or plasma samples^1,3,4^. Several studies have shown that saliva is suitable for both SARS-CoV2 viral RNA detection and serology measurements, but these measurements have not been made together using the same sample. In this study, we combined RT-qPCR with serology measurements on a small number of symptomatic COVID-19 patient saliva samples. We demonstrate that combining these measurements identifies SARS-CoV2 infection in patients that RT-qPCR alone misses, providing information that can help guide clinical decision making. This is particularly important in patients who present to the hospital with COVID-19 symptoms but have negative RT-qPCR results despite having been infected with SARS-CoV2 (for example because the immune system has already cleared the virus).

To maximize sensitivity of RNA detection, we developed and optimized a high-efficiency saliva RNA extraction protocol without the use of kits. We found our protocol yields much higher levels of RNA from saliva than a widely-used commercial isolation kit when performed manually. We also directly compared our RNA extraction method to several recently published no-extraction methods for SARS-CoV2 detection by RT-qPCR in saliva. Although these methods greatly increase ease and throughput of RT-qPCR testing, this comes at a cost of sensitivity. Our high-volume RNA extraction protocol allows for a 30-fold concentration of the saliva sample in addition to the inactivation and removal of enzymatic inhibitors present in saliva. In a comparison utilizing saliva spiked with virus, our high-volume RNA extraction protocol led to the detection of at least 56 times more viral RNA than the use of inactivated saliva as input.

To maximize the sensitivity of our serology measurements, we employed serology assays that we recently developed using Single Molecule Array (Simoa) technology^7^. The high sensitivity of these assays is particularly advantageous in saliva, where antibody levels are known to be low^19,30,34^. We have also demonstrated that serology measurements can be combined with direct, amplification-free detection of RNA in saliva samples. We envision that the sensitivity of the RNA assay can be further improved, for example, by adding Cas13a-based detection^49^. Future studies may also incorporate additional protein biomarkers in saliva to measure inflammation (cytokines, etc.) or other aspects of host response to increase the utility of multiplexed saliva diagnostics for COVID-19.

## Materials and methods

### Saliva samples

For protocol optimization and control experiments we used pooled human saliva in 1mL aliquots from BioIVT. Saliva was spiked with heat-inactivated SARS-CoV2 viral particles (ATCC) at the beginning of the protocol or with SARS-CoV2 synthetic RNA (Twist Bioscience) after Proteinase K treatment. COVID-19 positive and negative saliva samples were obtained from adult patients presenting to Massachusetts General Hospital (MGH). Seven samples were from patients who tested negative for SARS-CoV2 using RT-qPCR from nasopharyngeal (NP) swabs and 9 samples were from patients who tested positive for SARS-CoV2 using RT-qPCR from NP swabs. Pre-pandemic control samples were purchased from BioIVT. Samples were centrifuged at 13,150 × g at 4°C for 10 min. The supernatant was removed and used for RNA and antibody detection. All saliva samples were collected under approval of the Mass General Brigham Institutional Review Board for Human Subjects Research.

### RNA extraction using custom protocol

RNA extractions were done in 96-well LoBind PCR plates (Eppendorf) or 2000μL Deepwell plates (Eppendorf). Proteinase K and SDS were added to one volume of saliva (30μL or 300μL) to a final concentration of 0.5% SDS and 20x Proteinase K (New England Biolabs) and saliva was incubated at 65°C for 30 minutes. Two volumes of lysis buffer consisting of 6M Guanidinium Thiocyanate (Millipore Sigma) and 0.5% Triton X-100 (ThermoFisher Scientific) were then added and mixed by pipetting. Sera-Mag Carboxylate-Modified Magnetic Beads (Cytiva), used at 10μL per reaction, were washed twice in 1mL water and resuspended in an equivalent volume of lysis buffer. Beads in lysis buffer (10μL) were then added to each reaction. Three volumes of isopropyl alcohol (Millipore Sigma) were added to each reaction, and the samples were mixed well by pipetting up and down ten times. Samples were incubated for five minutes at room temperature to allow RNA to bind to beads, and then placed on a magnet. When solution cleared (approximately 2 minutes for small-volume extraction or 15-20 minutes for large-volume extraction), with the plate still on the magnet, supernatant was removed. When using low-volume (200μL) plates: beads were washed twice with 150μL of 70% ethanol. Ensuring that all of the ethanol is removed, RNA was then eluted using 10μL of nuclease-free water. When using high-volume (deep-well) plates: 150μL of 70% ethanol was added and beads were resuspended in 70% ethanol off of the magnet and transferred to a PCR plate. The PCR plate was then placed on magnet and supernatant was removed. Another 150μL of 70% ethanol was added to the deep-well plate to get any residual beads and transferred to beads on the PCR plate. One last 70% ethanol wash step was then performed on the PCR plate and all 70% ethanol removed. RNA was eluted using either 20μL (for recovery experiments) or 10μL (for comparison to other methods) of nuclease-free water.

### RNA extraction comparison

For all comparison experiments, pooled human saliva spiked in with heat-inactivated virus at 50 particles/μL was used. For the comparison of the custom protocol to the commercial kit, ThermoFisher Scientific MagMAX Viral/Pathogen Nucleic Acid Isolation was used according to manufacturer’s protocol with either 200μL input volume or 400μL input volume and 50μL elution volume. For the comparison to the no-extraction protocols, saliva with virus was transferred to PCR strip tubes for inactivation. For the 95°C protocol, saliva was heated at 95°C for 5 minutes. For the TCEP protocol, Tris(2-carboxyethyl)phosphine hydrochloride (Millipore Sigma) was dissolved in water, EDTA was added to a final concentration of 0.1M, and NaOH was added until the TCEP solution reached a pH of 8. This TCEP solution was added 1:100 to saliva and heated at 95°C for 5 minutes. For the SalivaDirect Proteinase K protocol, 50μL saliva was added to 6.25μL of Proteinase K, Molecular Biology Grade (New England Biolabs). The tube was then vortexed at 3200 RPM for 1 minute and heated at 95°C for 5 minutes. For all no-extraction methods, 8.2μL of inactivated saliva was used as input into the RT-qPCR.

### RT-qPCR

Viral RNA was quantified using Luna Universal Probe One-Step RT-qPCR kit (New England Biolabs) according to the manufacturer’s protocol in a CFX96 Real-Time Detection System (Bio-Rad). All RT-qPCR reactions were performed using CDC N1 primers^50^ (IDT), targeting the N-gene of SARS-CoV2, used at a final concentration of 0.4μM with the probe at 0.2μM. Sequences are: Forward: GACCCCAAAATCAGCGAAAT, Reverse: TCTGGTTACTGCCAGTTGAATCTG, Probe: FAM-ACCCCGCATTACGTTTGGTGGACC-BHQ1. For each reaction, 8.2μL (of purified RNA or inactivated saliva) input was used together with 10μL Luna Master Mix, 1μL Luna Enzyme Mix, and 0.8μL primer/probe mix. For each RNA extraction optimization or comparison experiment, an aliquot of SARS-CoV2 synthetic RNA (Twist Bioscience) or heat-inactivated SARS-CoV2 viral particles (ATCC) was run on the same RT-qPCR plate. Differences in RT-qPCR cycle threshold (Ct) values between the sample and in-plate control (ex. 10,000 viral particles) were calculated using the equation:

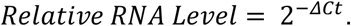

### Simoa Serology Measurements in Saliva

SARS-CoV2 serological measurements were performed as previously described^7^. The saliva samples were 100-fold diluted in ThermoFisher Scientific StartingBlock T20 Blocking Buffer (PBS) with 1X Halt Protease Inhibitor Cocktail (ThermoFisher Scientific) with EDTA. The Simoa assays were performed on the automated HD-X Analyzer (Quanterix). IgG, IgA and IgM against RBD, S1, spike, and nucleocapsid were measured in duplicate and the average AEB (average enzyme per bead) for each interaction was calculated.

### Direct SARS-CoV2 RNA detection with Simoa

A Simoa assay against SARS-CoV2 RNA was developed as described in detail in the Supplementary Information. Briefly, LNA capture probes were coupled to dye-encoded carboxyl-modified paramagnetic beads using EDC and Sulfo-NHS^48^. The beads were incubated with the sample at 60°C for 2 hours to allow hybridization of the LNA probes with the target RNA molecules. The beads were washed and incubated with biotinylated detector probes for 30 minutes. After additional washes, the beads were incubated with streptavidin-β-galactosidase (SβG) for 20 minutes. The beads were washed, resuspended in fluorogenic resorufin β-D-galactopyranoside (RGP) and loaded into a microwell array for imaging. The first two steps were performed offline in a 96-well plate, whereas the rest of the assay was performed on the HD-X Analyzer (Quanterix).

## Data Availability

Raw data is available upon request.

## Acknowledgments

The authors acknowledge funding from the Bill and Melinda Gates Foundation for this work. The MGH/MassCPR COVID biorepository was supported by a gift from Ms. Enid Schwartz, by the Mark and Lisa Schwartz Foundation, the Massachusetts Consortium for Pathogen Readiness and the Ragon Institute of MGH, MIT and Harvard.

