## Supplemental Information for "Ultrasensitive measurement of both SARS-CoV2 RNA and serology from saliva"

### Supplementary Information

Table of contents:


1. RNA extraction optimizations
2. Clinical characteristics of COVID-19+ saliva samples
3. Antibody profiles in saliva samples
4. Direct detection of SARS-CoV2 RNA using Simoa
5. Spike and recovery in control saliva samples

1. **<u>Figure S1: RNA Extraction Optimizations</u>**

A

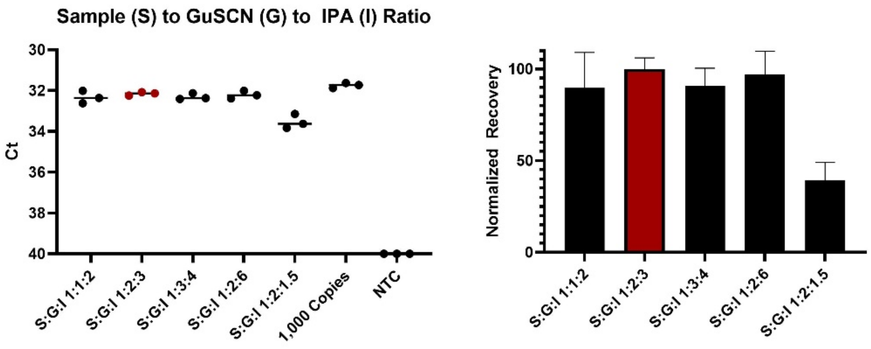


B.

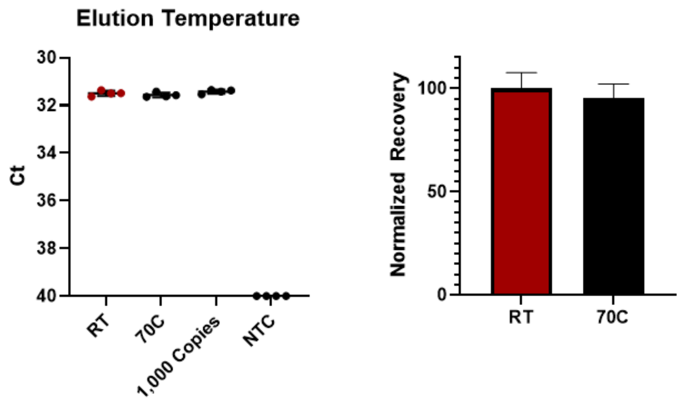


C

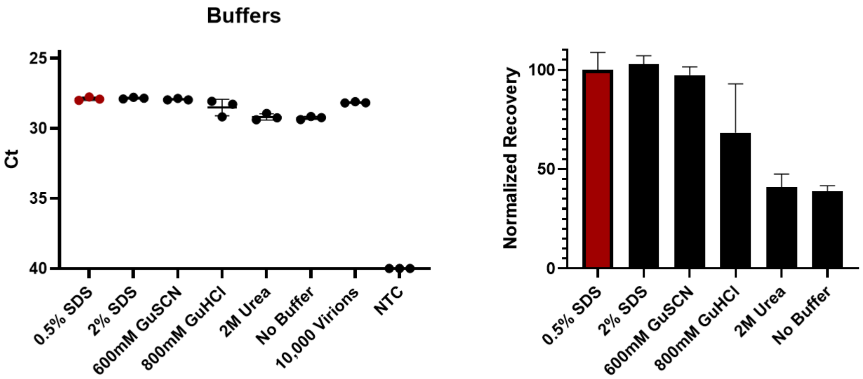


D

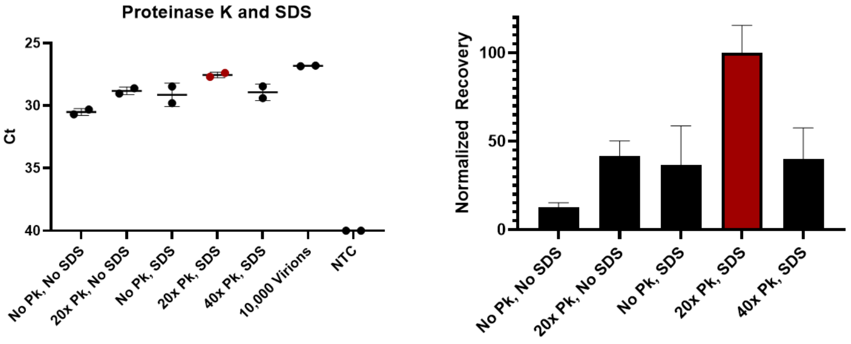

Supplementary Figure 1: RNA extraction optimizations.
Selected optimization experiments comparing extracted viral RNA to heat-inactivated virus or synthetic SARS-CoV2 RNA by RT-qPCR. NTC – No Template Control. Conditions were signal is undetected (NTC) are set to Ct = 40. A) Optimization of volumetric ratios of sample (PBS), guanidinium thiocyanate (GuSCN) and isopropyl alcohol (IPA) spiked with 1,000 copies of synthetic SARS-CoV2 RNA. B) Comparison of elution water temperatures from 30uL PBS spiked with 1,000 copies of synthetic SARS-CoV2 RNA. There was no significant difference (p = 0.4088, ns) between eluting RNA using room-temperature water versus eluting in water warmed to 70°C. C) Comparison of additives to enhance Proteinase K activity during the RNase inactivation step in 30µL saliva spiked with virus. D) Comparison of Proteinase K activity and SDS in 300µL of saliva spiked with virus. Recovery plots shown are normalized to final protocol (Custom Protocol: 0.5% SDS, Volumetric Ratios of Sample:GuSCN:IPA 1:2:3, 20x Proteinase K, RT elution, denoted by red color).

#### 2. Table S1: Clinical characteristics of saliva samples

| **Patient characteristics** | **NP PCR Positive (n = 9)** | **NP PCR Negative (n = 7)** |
|---|---|---|
| Age ($25^{th}$, $75^{th}$ quartile) | 38.9 (29, 47) | 30.7 (36, 27) |
| Male | 4 | 3 |
| Fever | 4 | 2 |
| Cough | 7 | 4 |
| SOB | 4 | 2 |
| Sore throat | 5 | 3 |
| Myalgia | 2 | NA |
| Obesity | 3 | 1 |
| Immunosuppression | NA | 1 |
| Lung disease | 2 | 1 |
| Heart disease | NA | NA |
| Required Ventilation | NA | NA |
| UTI | NA | 1 |
| Death | NA | NA |

#### 3. Figure S2: Antibody Profiles in COVID-19+ Saliva Samples

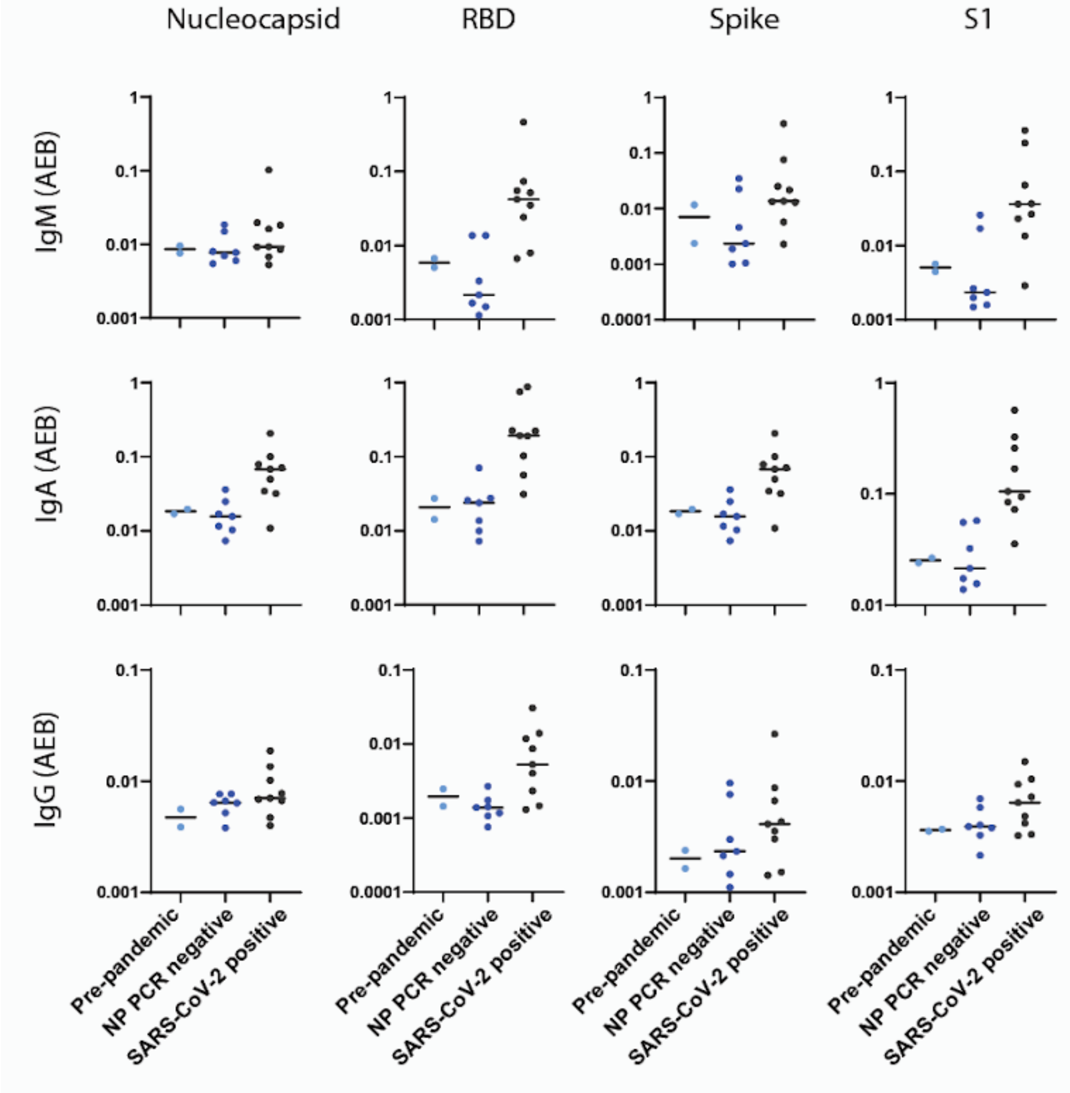


Supplementary Figure 2: Profiling antibodies in saliva samples.
Simoa serological assay results for IgG, IgM, and IgA against the four viral targets: spike, S1 subunit, RBD, and nucleocapsid for pre-pandemic samples (light blue, n = 2), NP RT-qPCR negative samples (blue, n = 7) and positive samples (dark blue, n = 9). Black lines indicate the median normalized AEB (average enzyme per bead) value of each population.

#### 4. Figure S3: Direct detection of SARS-CoV2 RNA in Simoa

Single Molecule Arrays (Simoa) have previously been adapted for direct detection of nucleic acids[1,2]. Here, we have developed a Simoa assay for the direct detection of SARS-CoV2 RNA. Paramagnetic beads were conjugated to 5' aminated LNA capture oligos using EDC and Sulfo-NHS, as previously described[2]. We based our capture and detector sequences on the EU SARS-CoV2 PCR primer set[3]. We used the EU Reverse (R) primer sequence, modified with LNA nucleotides and a poly-A extender, as a capture probe (SI Figure 3b). EU Forward (F) and Probe (P) sequences were biotinylated and used as detector probes. All capture and detector oligos were purchased from IDT.

Beads were prepared at concentrations of 240,000 beads/μL using an equal ratio of capture beads and three different dye encoded helper beads (Quanterix). 50μL of samples were added to a low-binding 96-well plate (Corning), and 5μL of well-mixed bead solution were added to each sample. To inactivate enzymatic RNases, 2μL of Proteinase K were added to each sample, and the plate was sealed, incubated at 60°C, and shaken at 1200 RPM for 30 minutes using an Eppendorf Thermomixer C. To inactivate cationic RNases, EDTA was added to the solution at a final concentration of 0.1mM. The plate was again sealed and incubated with shaking for 2 hours to allow bead-conjugated capture oligos to hybridize with the target RNA.

The plate was then moved to a magnet, and beads were washed twice with 150μl of 0.22μm filtered Hybridization Buffer (HB) (5x saline-sodium citrate [SSC] in diethyl pyrocarbonate [DEPC]-treated water). Detector solution was prepared by diluting EU F and EU P biotinylated probes to a final concentration of 25nM in HB. Beads were resuspended in 50μL of HB, and 5μL of Detector Solution was added to each sample. The plate was again incubated with shaking for 30 minutes at 60°C to allow detector probes to hybridize to the target RNA.

Beads were reconstituted in Dilution Buffer (1x sodium chloride–sodium phosphate–EDTA [SSPE] and 1.6% dextran sulfate in DEPC-treated water) and transferred onto a new 96-well plate (Quanterix). Streptavidin-β-galactosidase (SβG) Concentrate (Quanterix) was diluted to 50pM in SβG Diluent (Quanterix). The new plate and diluted SβG were loaded onto an HD-X analyzer (Quanterix). We used a 2-step assay and loaded Wash Buffer 1 in place of on-board beads and detectors. Beads were resuspended on-board in 100μL of SβG, incubated for 20 minutes (27 cadences), washed, resuspended with resorufin β-D-galactopyranoside (RGP) (Quanterix), and loaded into a microwell array for imaging.

All synthetic RNA spike in experiments used Twist Biosciences RNA standard. Sensitivity was greatly increased when RNA was fragmented. Thus, synthetic SARS-CoV2 RNA was fragmented using Zinc-based RNA fragmentation reagent (ThermoFisher Scientific) at 70°C for 5 minutes before being spiked into samples, according to the manufacturer's instructions.

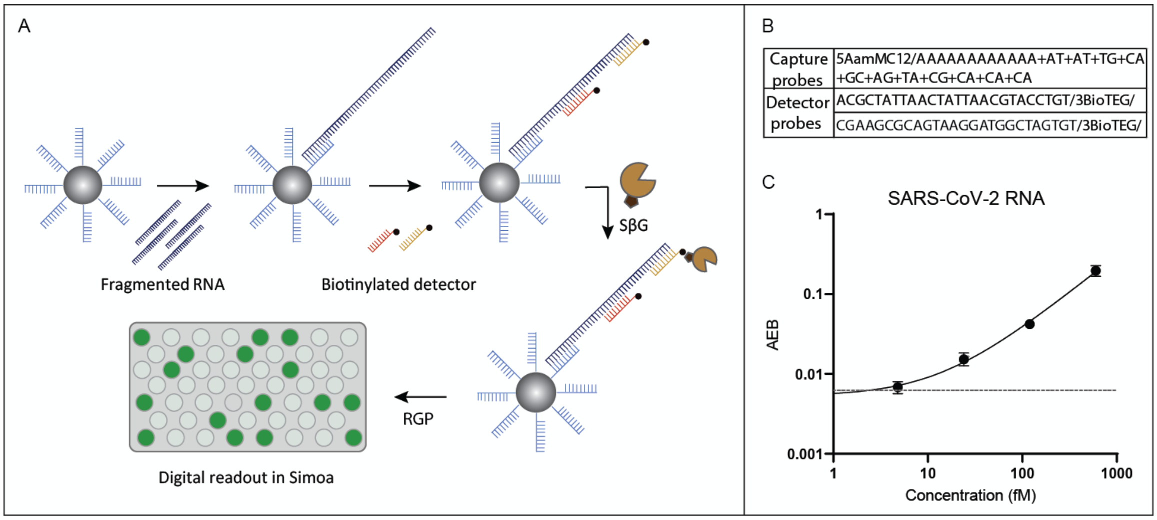


Supplementary Figure 3: Direct detection of SARS-CoV2 RNA in Simoa.

A) Schematic overview of Simoa RNA detection. Paramagnetic beads are conjugated with 5’ aminated capture oligos using EDC and Sulfo-NHS chemistry. Detection is done in three steps: 1) conjugated beads capture RNA, 2) biotinylated detector sequences hybridize with target RNA, 3) streptavidin beta-galactosidase binds to biotinylated detectors. The beads are finally resuspended with resorufin β-D-galactopyranoside (RGP) (Quanterix), and loaded into a microwell array for imaging. B) List of capture and detector sequences employed (5’ to 3’). C) A calibration curve of synthetic SARS-CoV2 RNA standard depicts a limit of detection (LOD) of 3.4fM RNA, or 102,000 copies of RNA, using our Simoa based SARS-CoV2 direct detection assay.

#### 5. Table S2: Spike and Recovery in Control Saliva Samples

| Analyte | Saliva Concentration | Water Concentration | Recovery (%) |
|---|---|---|---|
| SARS-CoV2 RNA | 222.073 fM | 243.641 fM | 91.1 |
| IgG Spike | 61.832 ng/mL | 51.950 ng/mL | 119.0 |
| IgG S1 | 69.757 ng/mL | 77.193 ng/mL | 90.4 |
| IgG RBD | 96.167 ng/mL | 89.335 ng/mL | 107.6 |
| IgM Spike | 70.250 ng/mL | 70.003 ng/mL | 100.4 |
| IgM S1 | 147.271 ng/mL | 155.652 ng/mL | 94.6 |
| IgM RBD | 272.338 ng/mL | 244.279 ng/mL | 111.5 |
| IgA Spike | 88.017 ng/mL | 59.119 ng/mL | 148.9 |
| IgA S1 | 54.185 ng/mL | 57.484 ng/mL | 94.260 |
| IgA RBD | 64.488 ng/mL | 58.529 ng/mL | 110.2 |

SARS-CoV2 negative control saliva and water samples were spiked with SARS-CoV2 RNA and anti-SARS-CoV2 IgG, IgM, and IgA antibodies against spike protein (CR3022). Samples were split, and RNA and antibody levels were detected simultaneously using the Simoa platform. For RNA, samples were diluted 4x into Hybridization Buffer (5x saline-sodium citrate [SSC] in diethyl pyrocarbonate [DEPC]-treated water). For serology, samples were diluted 100x into Sample Diluent (Quanterix). Recovery was measured as the percent of signal detected in saliva relative to water.
